# High levels of plasminogen activator inhibitor-1, tissue plasminogen activator and fibrinogen in patients with severe COVID-19

**DOI:** 10.1101/2020.12.29.20248869

**Authors:** David Cabrera-Garcia, Andrea Miltiades, Samantha Parsons, Katerina Elisman, Mohammad Taghi Mansouri, Gebhard Wagener, Neil L. Harrison

## Abstract

We measured plasma levels of fibrinogen, plasminogen, tissue plasminogen activator (t-PA) and plasminogen activation inhibitor 1 (PAI-1) in blood from 37 patients with severe coronavirus disease-19 (COVID-19) and 23 controls. PAI-1, t-PA and fibrinogen levels were significantly higher in the COVID-19 group. Increased levels of PAI-1 likely result in lower plasmin activity and hence decreased fibrinolysis. These observations provide a partial explanation for the fibrin-mediated increase in blood viscosity and hypercoagulability that has previously been observed in COVID-19. Our data suggest that t-PA administration may be problematic, but that other interventions designed to enhance fibrinolysis might prove useful in the treatment of the coagulopathy that is often associated with severe COVID-19.

## Introduction

COVID-19, the disease associated with infection by the severe acute respiratory syndrome coronavirus 2 (SARS-CoV-2), can cause upper respiratory tract (URT) infections that may progress to bilateral pneumonia and acute respiratory distress syndrome (ARDS) (Mehta *et al*., 2020; Zhang *et al*., 2020). Extra-pulmonary symptoms are also characteristic of severe COVID- 19, and these include systemic inflammation and cytokine release (del Valle *et al*., 2020), often combined with disorders of blood clotting and secondary manifestations of disease in the lung, kidney, brain, heart and liver (Gavriatopoulou *et al*., 2020).

Increased thrombosis and thrombotic complications have been widely associated with COVID- 19 (Bikdeli *et al*., 2020; Magro *et al*., 2020; Giannis *et al*., 2020; Klok *et al*., 2020; do Espírito Santo *et al*., 2020), and abnormal coagulation is correlated with COVID-19 severity (Tang *et al*., 2020*b*). Changes in conventional parameters such as international normalized ratio (INR), fibrinogen level and platelet counts may be useful for the rapid detection of hypocoagulation in COVID-19 patients (Tang *et al*., 2020*a*; Xiong *et al*., 2020), but do not fully explain the pathophysiology of hypercoagulability seen in some of these patients.

Our clinical observations of increased thromboembolic events in COVID-19 led us to use rotational thromboelastometry (ROTEM) to study the mechanisms of hypercoagulability in severe COVID-19. We found that there was a significant increase of fibrin-mediated clot viscosity (Roh *et al*., 2020), but the exact mechanism of this fibrin-mediated hypercoagulability remained elusive. Understanding of the pathogenesis of coagulopathy in COVID-19 is incomplete, although treatment with anticoagulants such as heparin (Tang *et al*., 2020*b*) and nadroparin (Klok *et al*., 2020) in severe COVID-19 infections has become widespread (Moores *et al*., 2020).

We hypothesized that coagulopathy associated with severe COVID-19 might be associated with deficits in fibrinolysis, as well as with alterations in the primary coagulation pathway, and we therefore designed a study based on patients admitted to intensive care with severe COVID-19 during the peak of the epidemic of COVID-19 in New York City in April and May 2020.

## Methods

### Human samples

We conducted a Columbia University IRB-approved (protocol #AAAS0172) prospective cohort study of 37 patients with severe COVID-19 and critical disease (respiratory distress, shock and/or multiorgan dysfunction) and 23 non-COVID-19 controls were enrolled. All patients received a standard reverse transcription-polymerase chain reaction (RT-PCR) test for SARS-CoV-2. All 37 of the COVID-19 patient group had clinical symptoms of severe infection requiring hospitalization, as well as a confirmatory RT-PCR test for SARS-CoV-2. The control patient group consisted of healthy volunteers and admitted surgical patients, with no clinical evidence of viral infection and a negative RT-PCR test within 5 days of sample collection. Volunteers were not remunerated for participating in the study and health status for the volunteers was determined by self-report. The study was performed at Columbia University Irving Medical Center in New York, N.Y., and COVID-19 patients are all from the “first wave” of the pandemic in April-May 2020. Blood was drawn into EDTA-containing tubes, rapidly centrifuged and plasma separated before being stored at -80 °C until analysis. No solvents or detergents were added to the plasma samples, which were handled in a biological safety cabinet under enhanced Bio Safety Level (BSL)-2 protocols.

### Enzyme-linked immunosorbent assay (ELISA)

ELISA kits with colorimetric output were used to determine the plasma concentrations of PAI-1 (ab184863, Abcam), fibrinogen (ab241383, Abcam), plasminogen (ab108893, Abcam) and tissue plasminogen activator (t-PA) (ab190812, Abcam) according to the manufacturer’s instructions. The plates were read at 450 nm using an automated microplate reader (Biotek Epoch). The mean intra-assay and inter-assay coefficient of variation for the assays were, respectively: PAI-1 (3%, 13%), Fibrinogen (2%, 9%), Plasminogen (4%, 6%) and t-PA (4%, 8%). We report for each subject the mean value from at least three replicate determinations, all with a coefficient of variation (CV) lower than 20%. The assay concentrations of each biomarker were determined using a standard curve, constructed and fit using a 4P logistic regression equation, and then values were corrected by the appropriate dilution factor in each case.

### Statistics

All statistical analyses were performed using Prism 8 (GraphPad Software, Inc). Comparisons between the COVID-19 and control groups were performed using the non-parametric Mann-Whitney U test, and correlations were assessed using Spearman’s correlation matrix. Blood levels of the biomarkers investigated are presented here as population medians, together with either the 95% confidence interval (CI) or the interquartile range (IQR). Additional detailed statistical information is presented in Figure Legends.

## Results

### Patient Data

**Controls:** 11 healthy volunteers and 12 same-day surgical patients were recruited as controls, provided informed consent by the patients or the surrogates and blood was drawn and processed as described above. The median age of all controls was 32 (IQR 29 – 57) years and 10 (43.5%) were female (**Table 1**). The median ages of healthy volunteers and in-hospital controls were 29 (IQR 26 – 32) years and 50 (IQR 32 – 64) years, respectively. Blood of the same-day surgical patients was drawn prior to the beginning of the surgical procedure.

**Table 1.** Demographics of control and COVID-19 groups.

| **Variable** | **Control** | **COVID-19** |
| --- | --- | --- |
| Subjects, n | 23 | 37 |
| Age, years, median (IQR) | 32 (29, 57) | 61 (52, 70) |
| Female, n (%) | 10 (43.5%) | 16 (42.1%) |

**COVID-19 patients:** 37 patients (21 males and 16 females, with a median age of 61, IQR 52 – 70, years) with severe COVID-19, all of whom required mechanical ventilation, were included in this study (**Table 1**). These patients, all severely ill from COVID-19 with multiple complications and a high rate of multi-organ dysfunction were cared for in a conventional intensive care unit (ICU) and a temporary ICU that was created within the operating rooms of CUIMC. Most of the samples were collected between 5 and 15 days following admission to the ICU (median 9; IQR 5 – 15 days, data from 28 of 37 patients).

The clinical characteristics of COVID-19 patients are listed in **Table 2**. The most common comorbidities were obesity (28 patients, 75.7%), hypertension (25 patients, 67.6%) and diabetes (18 patients, 48.6%). With regard to medications, 32 patients (86.5%) received hydroxychloroquine, 26 patients (70.3%) received azithromycin and only 3 patients (8.1%) received remdesivir. Steroids (dexamethasone, hydrocortisone or methylprednisolone) were administered to 20 of 37 patients, (54.0%) and 7 patients (18.9%) received systemic anticoagulation. Thrombotic events, including stroke, were observed in 16 of 37 patients (43.2%). 28 patients (75.7%) showed signs of acute kidney injury (AKI), with 18 at stage 3 (48.6%), and 14 patients (37.8%) required continuous renal replacement therapy (CRRT). During the hospitalization, 9 patients (24.3%) died and the remainder of the patients, 28 (75.7%), were discharged, mostly to acute rehabilitation. In **Table 3**, we report selected laboratory values for COVID-19 patients. As reported by others (Al-Samkari *et al*., 2020; Tang *et al*., 2020*b*; Fogarty *et al*., 2020), levels of D-dimer were significantly elevated [IQR 3.3 - 12.9 mg/L], as were levels of interleukin 6 (IL-6) [IQR 64 – 315 pg/mL].

**Table 2.**
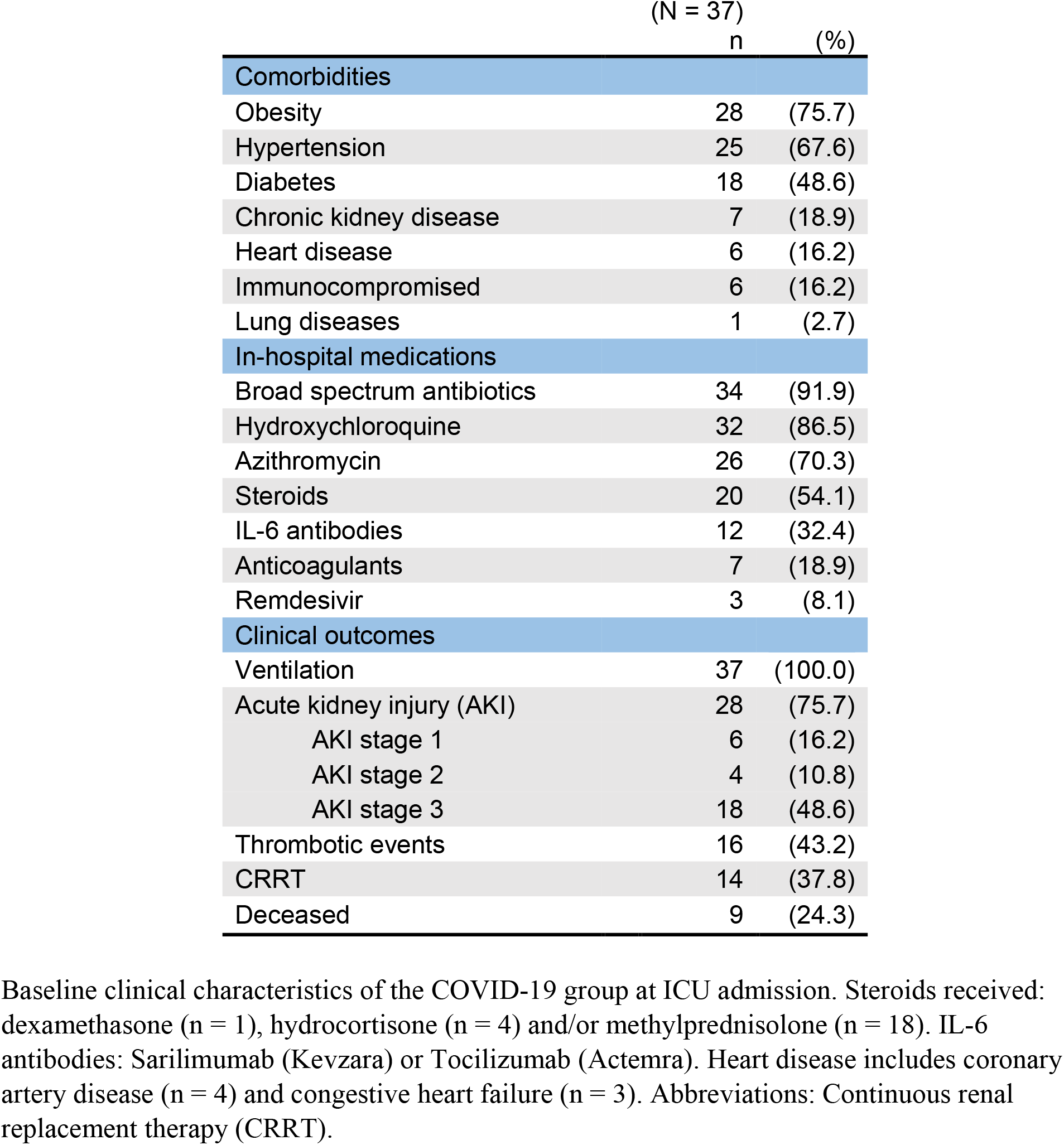
Clinical characteristics of patients with severe COVID-19.

**Table 3.** Laboratory results of COVID-19 patients.

|  | Reference values | COVID-19 (N = 37) |  |
| --- | --- | --- | --- |
|  |  | Median | IQR |
| **Coagulation markers** |  |  |  |
| INR | 0.8 – 1.2 | 1.2 | (1.2 – 1.3) |
| Prothrombin time (s) | 20 – 35 | 38.8 | (31.2 – 65.2) |
| D-dimer (µg/mL FEU) | < 0.8 | 6.5 | (3.3 – 12.9) |
| **Blood count** |  |  |  |
| Platelets (10<sup>9</sup>/L) | 150 – 350 | 277 | (197 – 352) |
| White blood cells (10<sup>9</sup>/L) | 4.5 – 11 | 11.6 | (7.9 – 16.2) |
| Hemoglobin (g/dL) | 12 -17 | 9.7 | (8.0 – 11.0) |
| **Markers of inflammation and organ injury** |  |  |  |
| IL-6 (pg/mL) | < 5 | 148 | (64 – 315) |
| Arterial lactate (mM) | 0.5 – 1 | 1.4 | (0.9 – 1.7) |
| Bilirubin (mg/dL) | < 0.3 | 0.5 | (0.4 – 0.7) |
| Serum creatinine (mg/dL) | 0.8 – 1.2 | 1.6 | (1.0 – 3.4) |

Values are reported as medians with IQR (interquartile range). Laboratory tests were conducted within one day of the date of collection of blood samples for ELISA assays. Abbreviations: International normalized ratio (INR), fibrinogen equivalent units (FEU).

#### Coagulation and Fibrinolysis Markers

Fibrinogen levels in patients with severe COVID-19 were significantly higher than in the control group (Controls: 2.5 [IQR 2.3 – 3.2] mg/mL, COVID-19: 6.6 [IQR 5.5 – 9.9] mg/mL, P < 0.0001) (**Fig. 1a**). No difference was found in plasminogen levels between controls and COVID- 19 patients (Controls: 163.2 [IQR 138.8 – 191.8] µg/mL, COVID-19: 148.3 [IQR 126.0 – 173.2] µg/mL, P = 0.3183) (**Fig. 1b**), but levels of t-PA were significantly higher in the COVID-19 group compared to controls (Controls: 3.9 [IQR 3.3 – 6.9] ng/mL, COVID-19: 23.0 [IQR 14.2 – 40.4] ng/mL, P < 0.0001) **Fig. 2a**).

**Figure 1.**
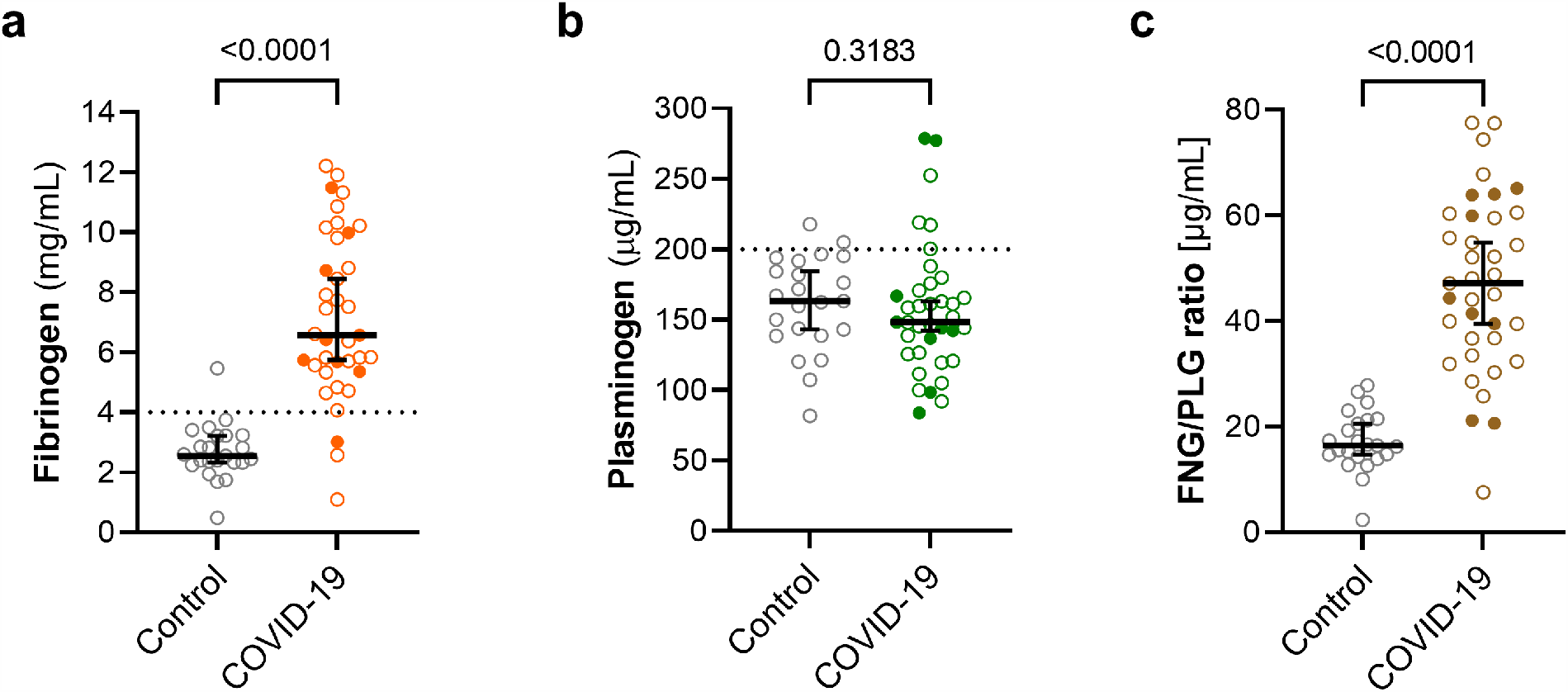
Plasma levels of fibrinogen are higher in COVID-19 patients. Fibrinogen (**a**) and plasminogen (**b**) were measured in plasma samples from severe COVID-19 patients (n = 37) and controls (n = 23). In addition, a ratio between fibrinogen (FNG, µg/mL) and plasminogen (PLG, µg/mL) was calculated (**c**). Graphs represent individual values with median and 95% confidence intervals are shown. Deceased patients are shown with filled symbols. Groups were compared using the Mann-Whitney U test (P-values are shown in each panel). The dotted lines in (**a**) and (**b**) represent the upper bound of the normal range for plasma levels of fibrinogen (4 mg/mL) and plasminogen (200 µg/mL).

**Figure 2.**
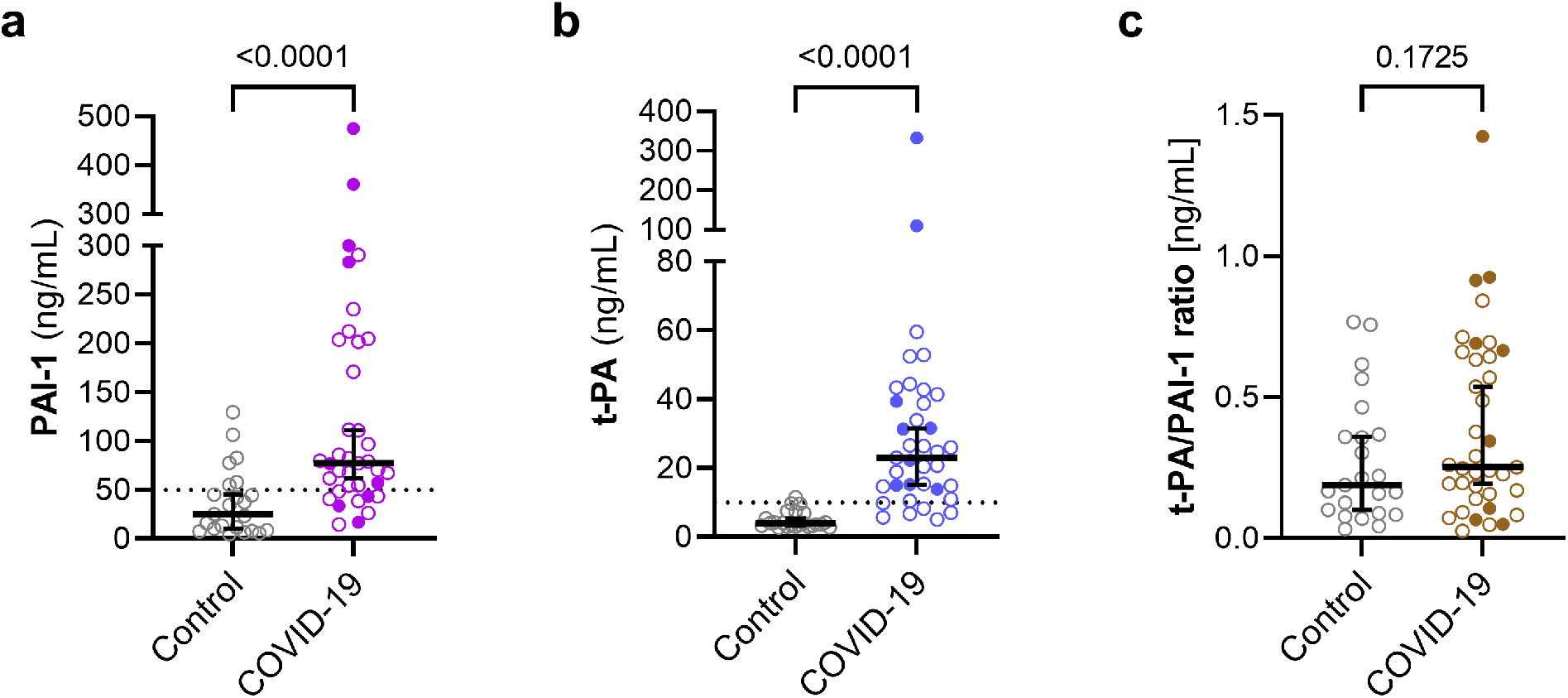
High levels of PAI-1 and t-PA are present in the plasma of COVID-19 patients. PAI-1 (**a**) and t-PA (**b**) were measured in plasma samples from severe COVID-19 patients (n = 37) and controls (n = 23). The three patients with the highest levels of PAI-1 and t-PA all expired (deceased patients shown with filled symbols). A ratio between t-PA (ng/mL) and PAI-1 (ng/mL) was calculated to express the relationship between these markers (**c**). The graphs display individual values, with the median shown by the horizontal bar. Deceased patients are shown with filled symbols. Groups were compared using the non-parametric Mann-Whitney U test (P- values are shown in each panel). Dotted lines in (**a**) and (**b**) represent the upper bound of the normal range for plasma levels of PAI-1 (50 ng/mL) and t-PA (10 ng/mL).

In addition, PAI-1 levels were elevated in COVID-19 patients (77.2 [IQR 51.4 – 202.6] ng/mL, P < 0.0001 compared to controls) (**Fig. 2b**). COVID-19 patients did not show a significant difference in the ratio (in ng/mL) between t-PA and its inhibitor PAI-1 when compared to controls (**Fig. 2c**).

Numerous COVID-19 patients showed levels of fibrinogen, t-PA and PAI-1 that were above the consensus normal range (**Fig. 1a, 2a, 2b**, dotted line).

Potential correlations between biomarkers were assessed using Spearman’s correlation matrix (**Fig. 3**). We observed a positive correlation between fibrinogen and plasminogen levels (r = 0.51; P = 0.0011) and a negative correlation of D-dimer with both fibrinogen (r = -0.39; P = 0.0156) and plasminogen (r = -0.51; P = 0.0013) (**Fig. 3**). Plasma levels of PAI-1 and t-PA were somewhat correlated (r = 0.31, P = 0.065) (**Fig. 3**). The number of white blood cells (WBC) was positively correlated with levels of t-PA (r = 0.35, P = 0.036) and D-dimer (r = 0.33; P = 0.045). IL-6 levels also correlate with higher D-dimer levels (r = 0.43; P = 0.0074), but only weakly with PAI-1 levels (r = 0.252; P = 0.1319) (**Fig. 3**).

**Figure 3.**
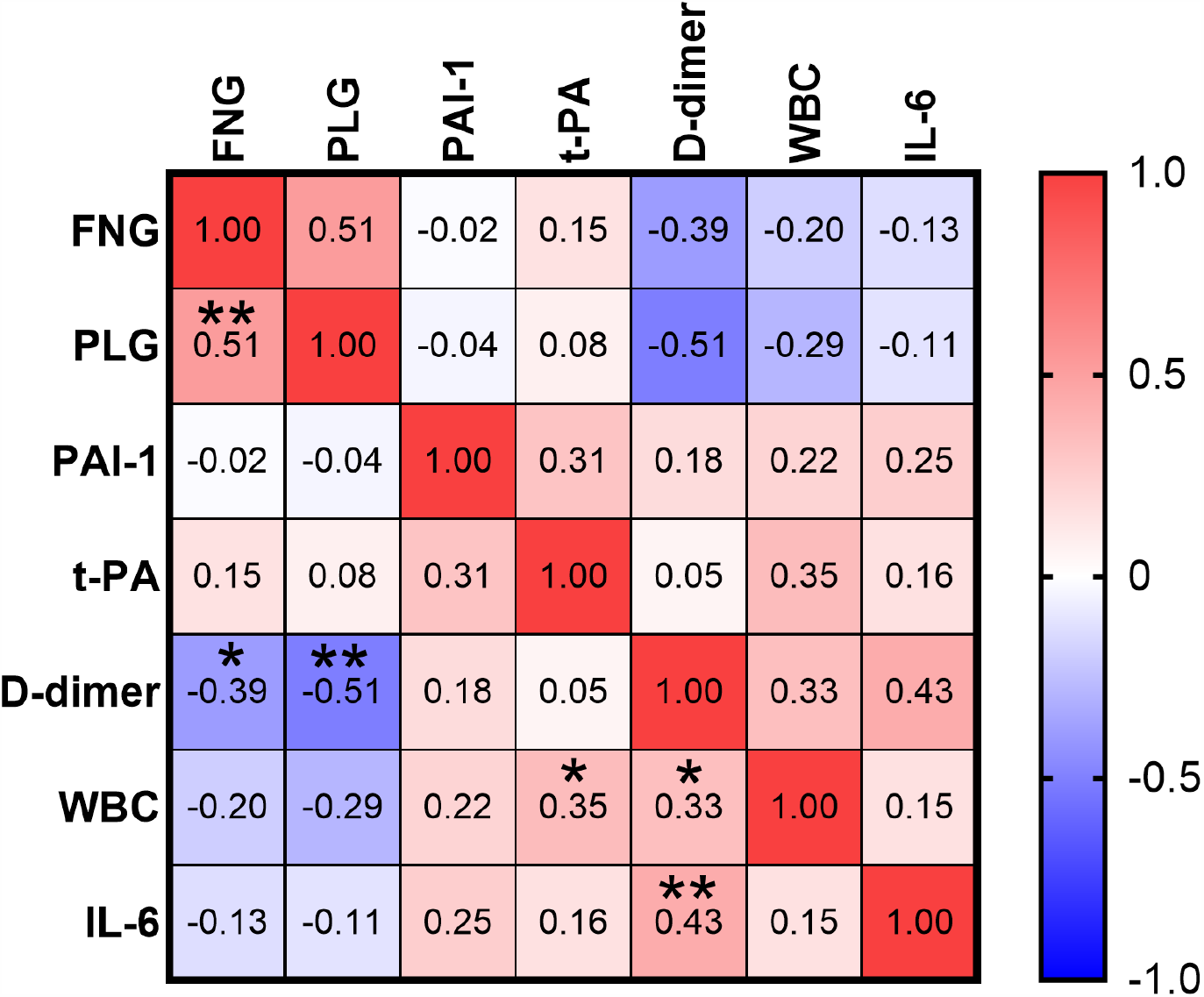
Correlation between markers of coagulation and fibrinolysis and other markers. Correlation matrix heat map. The values within the cells are Spearman correlation coefficients, and the depth of color corresponds to the strength of the correlation from very negative (dark blue) to very positive (dark red) correlation. Statistically significant correlations are noted as follows: * P < 0.05, ** P < 0.01. Abbreviations: WBC = white blood cell count, FNG = fibrinogen, PLG = plasminogen.

## Discussion

Our study demonstrated that patients with severe COVID-19 have elevated levels of PAI-1, tPA and fibrinogen but normal plasminogen levels compared to controls; evidence that could explain the fibrin mediated hypercoagulability seen in COVID-19. There is normally a homeostatic equilibrium between the process of coagulation, necessary for clot formation to prevent bleeding, and the process of fibrinolysis, which is required for normal wound healing (**Fig. 4**). Excessive fibrinolysis can result in abnormal bleeding, whereas a deficit in fibrinolysis can result in plaque formation, disseminated intravascular coagulopathy, stroke and thrombosis (Chapin & Hajjar, 2015). Some of the extensive changes in the biomarkers measured here might represent homeostatic adaptations to the imbalances created in COVID-19, whereas others may be primary alterations that result from the viral infection and/or the resulting immune response.

**Figure 4.**
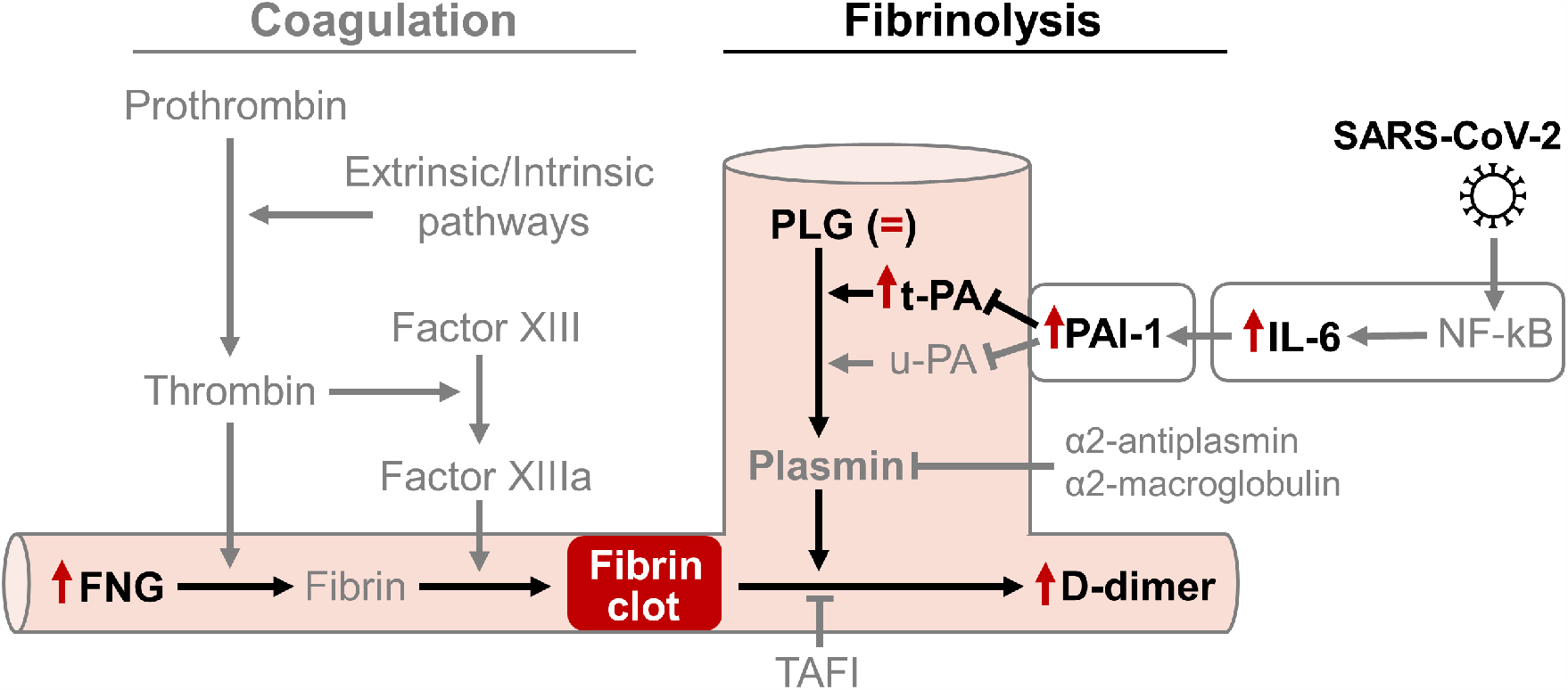
Diagram of the major components of the fibrinolytic and coagulation pathways, illustrating the role of the various biomarkers measured in this study. FNG (Fibrinogen), Plasminogen (PLG), t-PA (tissue plasminogen activator), u-PA (urine-type tissue plasminogen activator), IL-6 (interleukin 6), nuclear factor kappa-light-chain-enhancer of activated B cells (NF-κB), Thrombin-activatable fibrinolysis inhibitor (TAFI).

### Fibrinogen and plasminogen

Fibrinogen levels have also been shown to be elevated in other similar studies of COVID-19 patients (Nougier *et al*., 2020) and these findings and ours are quantitatively very similar. Increased fibrinogen levels are consistent with our prior observations on the important contribution of fibrinogen to the significant increase in clot viscosity in COVID-19 (Roh et al., 2020). It is not known whether the excess fibrinogen level represents a response to circulating cytokines or is a direct result of viral infection of the liver.

We found no significant change in plasminogen levels in COVID-19, although there was a positive correlation between levels of fibrinogen and plasminogen (**Fig. 3**). Both fibrinogen and plasminogen are synthesized in the liver (Raum *et al*., 1980; Tennent *et al*., 2007). Fibrinogen can be also synthesized extrahepatically in response to inflammation (Weisel, 2005), whereas plasminogen is thought to be almost exclusively produced in the liver (Raum *et al*., 1980). The higher ratio of fibrinogen to plasminogen (FNG/PLG ratio) in severe COVID-19 may therefore reflect the increased production of fibrinogen (but not plasminogen) at extrahepatic sites.

More surprising was a *negative* correlation between fibrinogen (and plasminogen) and levels of both D-dimer and IL-6 (**Fig. 3**). One interpretation of this result is that the very high levels of fibrinogen and plasminogen elevation represent the disease during the early part of the proposed “cytokine storm” and perhaps the peak of the coagulopathy, at which point the D-dimer breakdown product is still low. Extremes of elevated D-dimer may occur later on in the disease by which time fibrinogen and plasminogen have declined and the D-dimer waste product has accumulated, along with some organ damage. The inverse correlation between fibrinogen and IL-6 was also surprising, and might suggest that cytokines other than IL-6 are critical for the induction of the coagulopathy.

### PAI-1 and t-PA

Previous authors have proposed that there might be a “fibrinolysis shutdown” (Wright *et al*., 2020) or reduced fibrinolysis (Ji *et al*., 2020) in severe cases of COVID-19. The fibrinolytic system is controlled by a series of inhibitors that normally serve to limit the potential for bleeding, and one of the most important inhibitors of this system is the plasminogen activator inhibitor type 1 (PAI-1) (Cesari *et al*., 2010). Elevated levels of PAI-1 have been shown to increase the risk of atherothrombotic events (Vaughan, 2005; Baluta & Vintila, 2015). Over- expression of PAI-1 has been implicated in a variety of disease states, including atherosclerosis (Ploplis, 2011), obesity (Skurk & Hauner, 2004), and asthma (Ma *et al*., 2009), but the magnitude of such increases in PAI-1 are much lower than the very large increases reported here in COVID-19 patients. Similar observations reported by other groups in COVID-19 patients (Ranucci *et al*., 2020; Masi *et al*., 2020; Nougier *et al*., 2020; Zuo *et al*., 2020) are consistent with our findings.

The processing of plasminogen into plasmin, which is the enzyme responsible for the final degradation of fibrin clots, is mediated by t-PA (see **Fig. 4**). In agreement with others (Nougier *et al*., 2020; Zuo *et al*., 2020), we found an increase in t-PA levels in COVID-19 patients. This may lead to an increase in the risk of rare but serious bleeding in some patients (Zuo *et al*., 2020).

In COVID-19 patients, we found that the ratio between PAI-1 and t-PA was unaltered. It is unclear whether additional sites and mechanisms for the increased production of PAI-1 and t-PA may be involved, in addition to inflammation of endothelial cells. PAI-1, like t-PA, is normally produced and released from the vascular endothelium, but PAI-1 can also be produced by extravascular tissues, such as mast cells (Cho *et al*., 2000), and adipose tissue (Cesari *et al*., 2010), where its expression can be induced via activation of NF-κB (Okada et al 1998) by IL-1, IL-6, TNF α and a wide range of other pro-inflammatory mediators. These mediators are known as components of the proposed “cytokine release syndrome” that is the characteristic of many patients with severe COVID-19 (Del Valle et al., 2020). Certainly, one can say that markers such as IL-6 are now commonly used as predictors of COVID-19 severity (Del Valle *et al*., 2020), although this is not universally accepted (Mudd *et al*., 2020). *In vitro* data indicate that IL-6 induces the production of PAI-1 in cultured cells, while the inhibition of IL-6 signaling has been shown to reduce PAI-1 levels in COVID-19 patients (Kang *et al*., 2020).

We found that PAI-1 and t-PA were *positively* correlated with one another in our sample (**Fig. 3**). This is not unexpected, since both are produced in the vascular endothelium and might reflect a degree of tissue inflammation that results in excess release of these biomarkers. There was a *negative* correlation between t-PA (and PAI-1) and both D-dimer and IL-6. Again this can be interpreted in terms of different phases of the disease, with the extremes of PAI-1 and t-PA observed at perhaps the peak of the coagulopathy, and elevated D-dimer and IL-6 later on in the disease.

Coagulation disorders were also reported in the previous smaller epidemics associated with the related beta-coronaviruses, SARS-CoV-1 and Middle East Respiratory Syndrome coronavirus (MERS-CoV) (Giannis *et al*., 2020), and there is some evidence for PAI-1 involvement in the pathology of these infections. The SARS-CoV-1 N protein can induce the expression of PAI-1 in human peripheral lung epithelial (HPL1) cells *in vitro* (Zhao et al., 2008) via transforming growth factor-β (TGF- β), while the PAI-1 4G/5G gene polymorphism was found to be associated with the incidence of post-SARS osteonecrosis (Sun *et al*., 2014).

In comparative studies of patients with acute respiratory distress syndrome (ARDS) associated with COVID-19 and with other causes, high levels of PAI-1 and t-PA were reported in patients, irrespective of whether they had COVID-19 (Masi *et al*., 2020). In other studies, the specific clinical characteristics and biochemical mechanisms involved in COVID-19 appear to be somewhat distinct from what is commonly described in sepsis (Iba *et al*., 2020).

It remains unclear whether the coagulopathy observed in COVID-19 is unique, or may be similar to what is described for other forms of systemic coagulopathy, and this is clearly an important topic for future clinical investigations, in order to resolve these questions by the study of the biomarkers described here in populations of septic patients, with and without COVID-19.

### Implications for therapeutic intervention and patient management

#### t-PA use may be contraindicated in some patients with COVID-19

From the early stages of the COVID-19 pandemic, it has been clear that thrombosis and other complications associated with coagulopathy are a feature of the disease, and attempts to rectify the situation by stimulating fibrinolysis have been reported and discussed widely (Bornstein & Páramo, 2020), with the initial focus exclusively on t-PA. Most of the early t-PA studies lack a control arm, and are thus difficult to interpret, even though clinical trials of this intervention appear to be underway (Moore *et al*., 2020). In addition, as discussed by Zuo *et al*. (2020), the use of t-PA in severe COVID-19 appears problematic for many reasons, with which we concur.

Firstly, some COVID-19 patients appear to have levels of t-PA that are already extremely elevated, as shown here and by others. This suggests that measurement and monitoring of t-PA might help to detect rare but fatal bleeding events that have been documented by others (Zuo *et al*., 2020). Secondly, patients with extremely high levels of t-PA may already be at risk for rare but serious bleeding (Zuo *et al*., 2020). In our study, two patients with the highest levels of t-PA did not survive. Such patients would be detectable soon after admission by determining t-PA and PAI-1 and calculating the ratio between t-PA and PAI-1, and this could be investigated in larger studies. Thirdly, t-PA administered exogenously, is known to have a short half-life (Verstraete *et al*., 1985), and indeed the administration of this drug to typical COVID-19 patients with severe disease would likely result in rapid inactivation, due to the very high levels of ambient PAI-1 that are already present in these individuals.

Given the data here and elsewhere showing the presence of very high levels of PAI-1 and of t- PA itself in COVID-19 patients, skepticism seems warranted concerning the clinical utility of t- PA, especially in view of the dangers of “unexpected” bleeding and the poor prognosis associated with very high levels of t-PA (Zuo *et al*., 2020). Streptokinase might be a useful alternative treatment for COVID-19 coagulopathy, since it activates plasminogen by a distinct mechanism (Young *et al*., 1998) and there is evidence that streptokinase administered via a nebulizer can be effective in coagulopathy in ARDS resulting from other diseases (Abdelaal Ahmed Mahmoud *et al*., 2020).

#### PAI-1 antagonists may be useful in COVID-19 coagulopathy

A primary excess of PAI-1 may contribute to an imbalance between coagulation and fibrinolysis, resulting in the widespread and persistent coagulopathy seen in some COVID-19 patients. Inhibitors of PAI-1 such as tiplaxtinin (PAI-039) already exist (Elokdah *et al*., 2004; Izuhara *et al*., 2008; Boe *et al*., 2013) and these could perhaps be considered as experimental therapeutics for emergency use in COVID-19, supplementing the use of anticoagulants and corticosteroids.

PAI-1 antagonists may synergize with these drugs in the treatment of COVID-19 coagulopathy, perhaps reducing the doses of anticoagulant and steroids required, and thereby minimizing unwanted side effects. Monitoring of PAI-1 and t-PA levels and other routine coagulation parameters would in any case be required to assure safe use of any of these drugs, which should be restricted to patients within the ICU, to minimize the possibility of bleeding due to the elevated levels of t-PA that are often present.

## Conclusions

Our study has some obvious limitations, including a relatively small sample size and the use of a single time point for measurement of the biomarkers. Nevertheless, the group of severe COVID- 19 patients studied here can be considered as a representative sample from the peak of the first wave of COVID-19. The reproducible nature of these findings in a variety of different medical centers (Jin *et al*., 2020; Nougier *et al*., 2020; Zuo *et al*., 2020) supports the idea that changes in PAI-1 and fibrinogen, in particular, represent robust and universal biomarkers for COVID-19 that can be assessed in the ICU setting. Given the small size of our study, it remains unclear whether the changes in these biomarkers, and in the abnormal coagulation measured by ROTEM (Roh *et al*., 2020) is related to the incidence of thrombosis, myocardial infarction, stroke, renal failure and a variety of long-term complications in these patients. Determining these clinical relationships will require the extension of the current investigations into much larger scale laboratory and outcome studies.

## Data Availability

All data are available from the corresponding authors upon reasonable request.

## ACKNOWLEDGEMENTS

This work was supported solely by the Department of Anesthesiology at CUIMC. We thank C.W. Emala for helpful discussions and advice on ELISA assays. We thank Peter Yim for helpful discussion. We are grateful to M. Shelanski and A. Brambrink for logistical support of our laboratories during the pandemic. We are deeply grateful to the patients and their families for their participation in this study

## AUTHOR CONTRIBUTIONS

NLH, GW and DCG conceived the study and designed the experiments. AM, KE and GW recruited and consented patients. SP, KE and AM retrieved patient data. GW, KE, DCG, NLH and MM processed samples, and DCG performed the ELISA experiments and data analysis. All authors contributed to data analysis and interpretation. NLH, DCG and GW wrote the first draft of the manuscript and all authors commented on and edited the manuscript.

